# Interplay between climate, childhood mixing, and population-level susceptibility explains a sudden shift in RSV seasonality in Japan

**DOI:** 10.1101/2025.03.02.25323095

**Authors:** Sang Woo Park, Inga Holmdahl, Emily Howerton, Wenchang Yang, Rachel E. Baker, Gabriel A. Vecchi, Sarah Cobey, C. Jessica E. Metcalf, Bryan T. Grenfell

## Abstract

Titrating the relative importance of endogenous and exogenous drivers for dynamical transitions in host-pathogen systems remains an important research frontier towards predicting future outbreaks and making public health decisions. In Japan, respiratory syncytial virus (RSV), a major childhood respiratory pathogen, displayed a sudden, dramatic shift in outbreak seasonality (from winter to fall) in 2016. This shift was not observed in any other countries. We use mathematical models to identify processes that could lead to this outcome. In line with previous analyses, we identify a robust quadratic relationship between mean specific humidity and transmission, with minimum transmission occurring at intermediate humidity. This drives semiannual patterns of seasonal transmission rates that peak in summer and winter. Under this transmission regime, a subtle increase in population-level susceptibility can cause a sudden shift in seasonality, where the degree of shift is primarily determined by the interval between the two peaks of seasonal transmission rate. We hypothesize that an increase in children attending childcare facilities may have contributed to the increase in susceptibility through increased contact rates with susceptible hosts. Our analysis underscores the power of studying infectious disease dynamics to titrate the roles of underlying drivers of dynamical transitions in ecology.

## Introduction

Characterizing the drivers of dynamical transitions is a fundamental challenge in ecology (Earn et al., 2000; Hastings, 2004; Hastings et al., 2018). However, time series data from ecological systems are rare, and, where they do exist, sparse; reducing our ability to tease apart the relative roles of endogenous (e.g., density-dependent responses) and exogenous (e.g., climate variables) factors in driving dynamical transitions (Hunter and Price, 1998; Lundberg et al., 2000; Hernandez Plaza et al., 2012). There is one important exception: detailed spatiotemporal surveillance data are available for many epidemiological systems, providing a unique platform for answering broader questions in ecology and population biology (Levin et al., 1997; Anderson and May, 1991; Grenfell et al., 2001; He et al., 2010).

Respiratory syncytial virus (RSV) is a common childhood respiratory pathogen that infects nearly all children by the age of two, and is also an important risk factor for asthma and allergy development (Sigurs et al., 1995, 2010; Edwards et al., 2012). RSV outbreaks typically exhibit annual or biennial patterns with relatively consistent seasonal incidence across years in many countries, including Canada (Paramo et al., 2023), Korea (Kim et al., 2020), and the US (Pitzer et al., 2015; Baker et al., 2019). Previous studies showed that climate-driven transmission plays a major role in driving RSV epidemic dynamics (Pitzer et al., 2015; Baker et al., 2019). In particular, Baker et al. (2019) demonstrated a quadratic relationship between specific humidity and RSV transmission with minimum transmission occurring at intermediate humidity.

RSV in Japan presents a unique case study relative to other countries: In contrast to stable seasonal incidence generally observed, a sudden, dramatic transition from winter to fall RSV outbreaks was observed in Japan in 2016 (Miyama et al., 2021; Wagatsuma et al., 2021). To our knowledge, this change was not observed in other countries, including Australia (Nazareno et al., 2022), Canada (Public Health Agency of Canada, 2024), China (Luo et al., 2022; Li et al., 2023, 2024), Korea (Kim et al., 2023), and the US (Rose, 2018; Hansen et al., 2022). Wagatsuma et al. (2021) hypothesized that changes in climate and an increase in inbound overseas travelers may be jointly responsible for this shift in seasonality. However, their conclusion relied on correlational analyses, and little ecological support was provided for their proposed mechanism. Understanding the sudden shift in RSV seasonality is a necessary step for predicting future RSV outbreaks, as well as for timely deployment of monoclonal antibodies and vaccination (Mazur et al., 2023).

Here, we analyzed the time series of RSV cases from Japan (Fig. 1) to identify the drivers of a sudden shift in seasonality between 2016 and 2017. We combined a parsimonious model of disease transmission with a Bayesian statistical framework to infer RSV transmission dynamics across different islands. We used inferred transmission patterns to explore how changes in susceptibility can lead to a sudden shift in seasonality. Our analysis offers novel insights into drivers of dynamical transition in seasonal respiratory epidemics.

**Figure 1:**
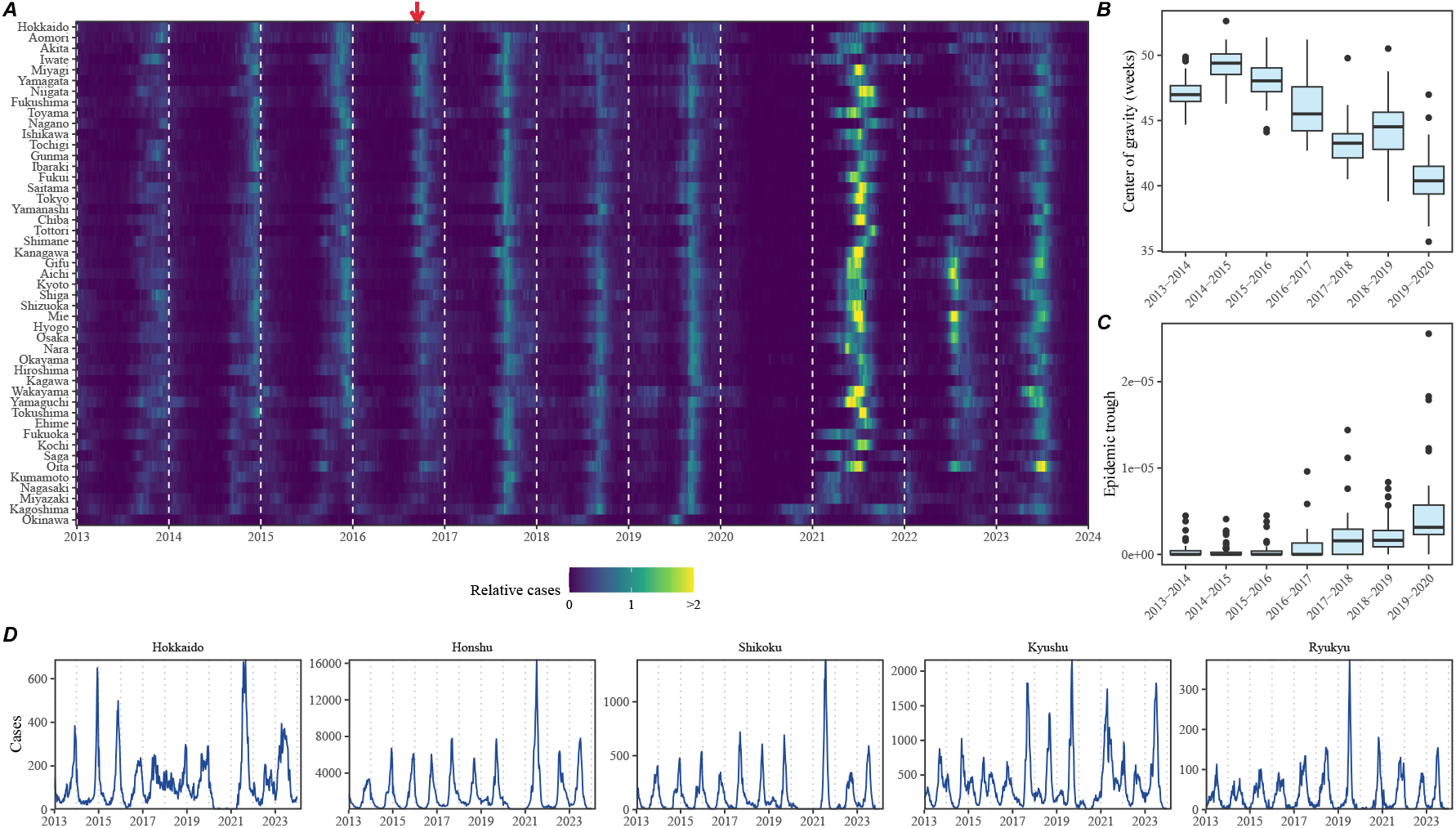
Observed changes in RSV outbreak dynamics in Japan. (A) Relative RSV cases across 47 prefectures in Japan between 2013 and 2024. Relative cases are calculated by dividing the raw cases by the pre-pandemic maximum. The red arrow indicates when the shift in seasonality occurred. (B) Estimates of center of gravity (i.e., the mean timing of an epidemic) across 46 prefectures, excluding Okinawa. (C) Estimates of the epidemic trough (i.e., the minimum number of cases in a season divided by the population size) across 47 prefectures. (D) Time series of RSV cases across 5 major islands.

## Results

### Observed dynamics in RSV outbreaks

A sudden change in RSV seasonality from winter to fall outbreaks was observed in nearly all prefectures between 2016 and 2017 (Fig. 1A). To quantify changes in seasonality, we calculated the center of gravity (i.e., the mean timing of an epidemic) for each outbreak season at every prefecture and found a consistent decrease in the center of gravity (Fig. 1B; Supplementary Figure S1; Methods). We also found that these changes were associated with the inter-epidemic troughs becoming shallower (i.e., bigger minimum) (Fig. 1C).

We found considerable heterogeneity in the observed outbreak dynamics across the major islands, especially following the changes in seasonality (Fig. 1D; see Supplementary Figure S2 for the map of Japan). For example, annual RSV outbreaks in Hokkaido island became more persistent, causing high numbers of cases throughout the year. Semiannual RSV outbreaks in Shikoku and Kyushu islands became more annual with higher intensity, leading to sharper epidemics. Finally, in contrast to all other islands, RSV outbreaks in Ryukyu island exhibited summer outbreaks, which also became more intense leading up to 2020. We note that Hokkaido and Ryukyu each consist of one prefecture, Hokkaido and Okinawa, respectively, which correspond to top and bottom rows in Fig. 1A; therefore, the observed RSV dynamics in Honshu, Shikoku, and Kyushu islands represent the majority of RSV transmission in Japan.

### A parsimonious model for RSV epidemics

We first began by asking whether a simple Susceptible-Infected-Recovered-Susceptible (SIRS) model can capture the observed RSV outbreak dynamics in Japan, including the sudden change in outbreak seasonality. The SIRS model is the simplest dynamical model that allows for the possibility of immune waning and therefore represents one of the most parsimonious models for explaining outbreak dynamics of respiratory infections. Here, we extended the standard SIRS model such that we could simultaneously estimate periodic seasonal transmission rates and non-periodic changes in transmission due to NPI measures that were implemented to prevent COVID-19 (Materials and methods).

Accounting for flexible changes in seasonal transmission rates that deviate from standard sinusoidal shapes allowed the SIRS model to reproduce the observed dynamics across all five islands, including changes in seasonality that occurred during 2016–2017 as well as post-pandemic changes in outbreak patterns (Fig. 2A). In contrast to most seasonal respiratory pathogens, which exhibit an annual cycle in transmission pattern, we estimated semiannual peaks in transmission rates in four islands (Honshu, Shikoku, Kyushu, and Ryukyu), which were explained by the quadratic effects of specific humidity (Fig. 2B): in line with Baker et al. (2019), we estimated that transmission would peak at a low and high mean specific humidity(Fig. 2C). Interestingly, we found that minimum RSV transmission occurred at a much higher mean specific humidity in Ryukyu island than in other three islands (Fig. 2C). We did not find semiannual transmission rate patterns or quadratic humidity-transmission relationship for Hokkaido island (Fig. 2B–C); instead, we found that transmission decreased at very low specific humidity (*<* 5 g*/*kg). Combining transmission rate estimates from all five islands still gave a quadratic humidity-transmission relationship for specific humidity between *approx*5 g*/*kg and ≈ 18 g*/*kg, but the joint relationship poorly captured the humidity-transmission relationship in the Ryukyu island (Supplementary Figure S3).

**Figure 2:**
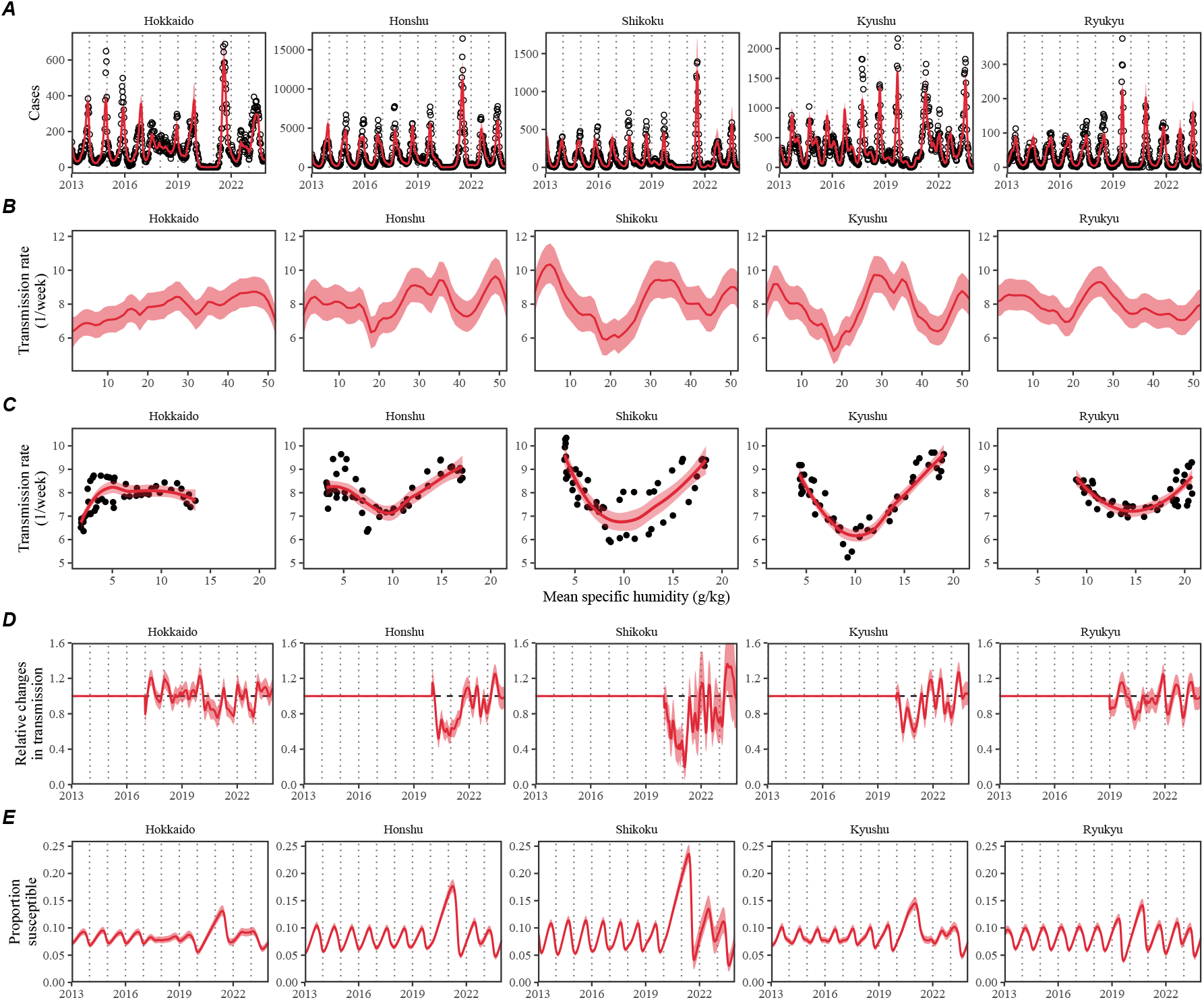
Summary of SIRS model fits to RSV outbreaks across major islands in Japan. (A) Comparisons of observed cases (points) across the five major islands and fitted epidemic trajectories (red lines). (B) Estimated periodic seasonal transmission rates. (C) Relationship between the estimated periodic seasonal transmission rates and mean specific humidity. Points represent seasonal transmission rate estimates across 52 weeks versus average humidity across 2013–2020. Lines represent the corresponding locally estimated scatterplot smoothing (LOESS) estimates. (D) Estimated relative changes in transmission, capturing the impact of NPI measures. (E) Estimated proportion of the susceptible pool. Lines represent the estimated median of the posterior distribution. Shaded regions represent the 95% credible intervals from the posterior distribution.

Across all five islands, we estimated large reductions in transmission during 2020 (Fig. 2D); however, there was large heterogeneity in the overall shape of the estimated NPI effects as well as the degree of transmission reduction. The reduction in transmission rates caused an increase in the susceptible pool (Fig. 2E), which allowed a large outbreak when NPIs were lifted (Fig. 2A). This observation is also consistent with Baker et al. (2019) who predicted that an accumulation of the susceptible host population during the NPI period will eventually lead to a large outbreak.

### Mechanisms for sudden changes in seasonality

Since the mechanistic SIRS model was able to accurately capture the observed transition in seasonality, we posit that it captures the relevant mechanisms for driving this pattern. Thus, we should be able to use the inferred dynamics to further tease apart the mechanisms underlying sudden changes in seasonality of the RSV outbreaks. To do so, we first began by evaluating the changes in the proportion of susceptible and infected individuals at the beginning of the season between 2013 and 2019. We focused on three main islands that exhibited clear changes in seasonality: Honshu, Shikoku, and Kyushu islands.

Across three main islands, we found a consistent increase in the proportion of susceptible and infected individuals at the beginning of each season between 2013 and 2019 (Fig. 3A). These changes also corresponded with a decrease in center of gravity (Fig. 3A). A more detailed comparison of epidemic trajectories illustrated that an increase in the susceptible pool at the beginning of the season can drive a sudden shift in seasonality (Fig. 3B). The semiannual pattern in transmission (i.e., two peaks in transmission rates within a year) allows this transition, where a faster epidemic growth (from higher susceptible pool) drives a faster depletion of susceptibles and causes the epidemic to transition from a later peak to an earlier peak.

**Figure 3:**
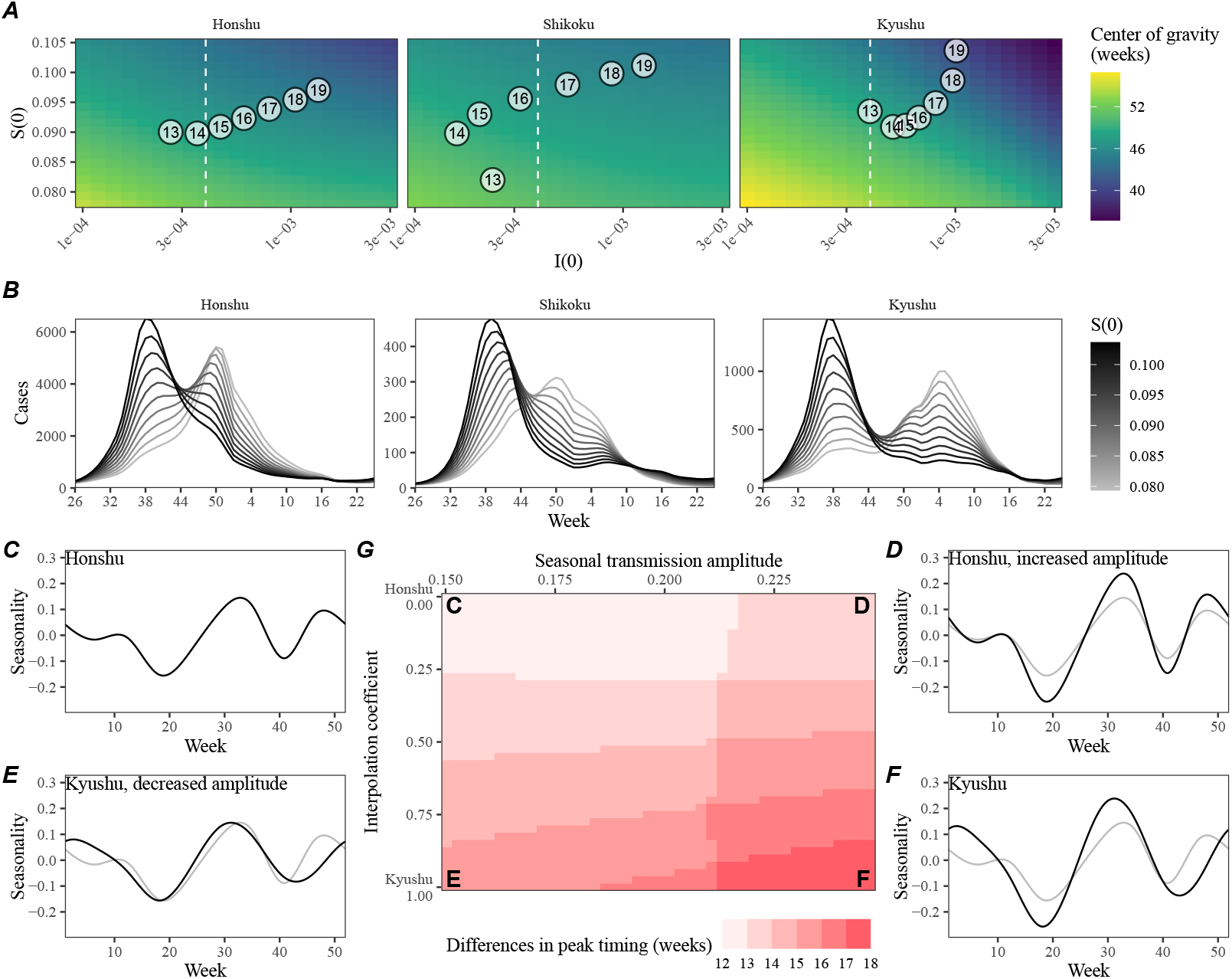
An increase in the susceptible pool explains sudden changes in seasonality. (A) Predicted effects of the proportion of infected *i*(0) and susceptible *S*(0) at the beginning of season on center of gravity. Points represent the estimated values for *i*(0) and *s*(0) between 2013 and 2019, showing the last two digits of a given year. The white vertical dashed line represents the *i*(0) value used for simulating epidemic dynamics in panel B. (B) Changes in epidemic trajectories caused by an increase in the susceptible proportion at the beginning of season for a fixed value of *i*(0). (C–F) Comparisons of interpolated transmission rates used for simulating the SIRS model, corresponding to each corner in Panel G. Black lines represent the transmission rates used for simulations. Gray lines represent the estimated transmission rates the Honshu island as a visual reference. (C) The estimated transmission rates for the Honshu island. (D) The resulting transmission rates for the Honshu island with equal amplitude as the Kyushu island. (E) The resulting transmission rates for the Kyushu island with equal amplitude as the Honshu island. (F) The estimated transmission rates for the Kyushu island. (G) Differences in peak epidemic timing when we increase the the susceptible proportion at the beginning of season from 7.8% to 10.5% using transmission patterns that interpolate the estimates for Honshu and Kyushu islands.

To understand why we observe a bigger shift in seasonal outbreak patterns in Kyushu island than in Honshu and Shikoku islands, we characterized how differences in the seasonal transmission patterns affects the difference in peak epidemic timing associated with an increase in population-level susceptibility (Fig. 3C–G). As a reference, we began with smoothed transmission rate estimates for Honshu (Fig. 3C) and Shikoku (Fig. 3F) islands and explore intermediate transmission patterns that interpolate two transmission patterns. Specifically, the transmission pattern in Kyushu exhibited a bigger amplitude and a wider trough between two transmission peaks. These differences were explored by varying the amplitude of the seasonal transmission rate (*x*-axis on Fig. 3G) and the degree of interpolation (*y*-axis on Fig. 3G), where the interpolation coefficients of 0 and 1 correspond to the seasonality in Honshu and Kyushu islands, respectively. Simulating the SIRS model using interpolated seasonal transmission rates showed that a large seasonal amplitude and wider trough cause larger changes in the timing of epidemic peak (Fig. 3G).

We did not observe a shift in RSV seasonality in Ryukyu island (Fig. 1E) despite the inferred quadratic humidity-transmission relationship (Fig. **??**B). In Supplementary Materials, we show that there was limited variation in population-level susceptibility between 2014–2019 in Ryukyu island, explaining the lack of change in RSV seasonality (Supplementary Figure S4A).

### Mechanisms for an increase in population-level susceptibility

So what mechanisms caused the population-level susceptibility to increase over time in Japan? We hypothesized that the Comprehensive Support System for Children and Childcare, which was enacted in 2012 and launched in 2015, brought more children into the daycare system and thus contributed to an increase in population-level susceptibility. This hypothesis builds on the work of DeHaan et al. (2024) who previously suggested that increase in childcare attendance may have contributed to a large change in the seasonality of Kawasaki disease in Japan in the mid-2010s. An expansion of childcare facilities would increase contact rates among children, which in turn would increase the probability of exposure and therefore the effective susceptibility against RSV infections. To quantify the potential impact of this program, we compared the number of children attending childcare facilities in Japan since 2013 and compared them with our estimates of susceptible proportion (Fig. 4). Overall, we found consistent patterns of increase in childcare attendance and strong correlations with the estimated susceptible proportion: 0.977 (95% CI: 0.847–0.997) in Honshu island, 0.943 (95% CI: 0.653–0.992) in Shikoku island, and 0.802 (95% CI: 0.124–0.970) in Kyushu island. The lack of island-level data did not allow us to test whether constant susceptibility estimated for the Ryukyu island is correlated with a lack of increase in childcare facilities in Ryukyu island; however, counterfactual simulations show that an increase in susceptibility would have been able to shift the RSV outbreaks to spring in Ryukyu island, consistent with predictions for other islands based on the quadratic humidity-transmission relationship (Supplementary Figure S4B).

**Figure 4:**
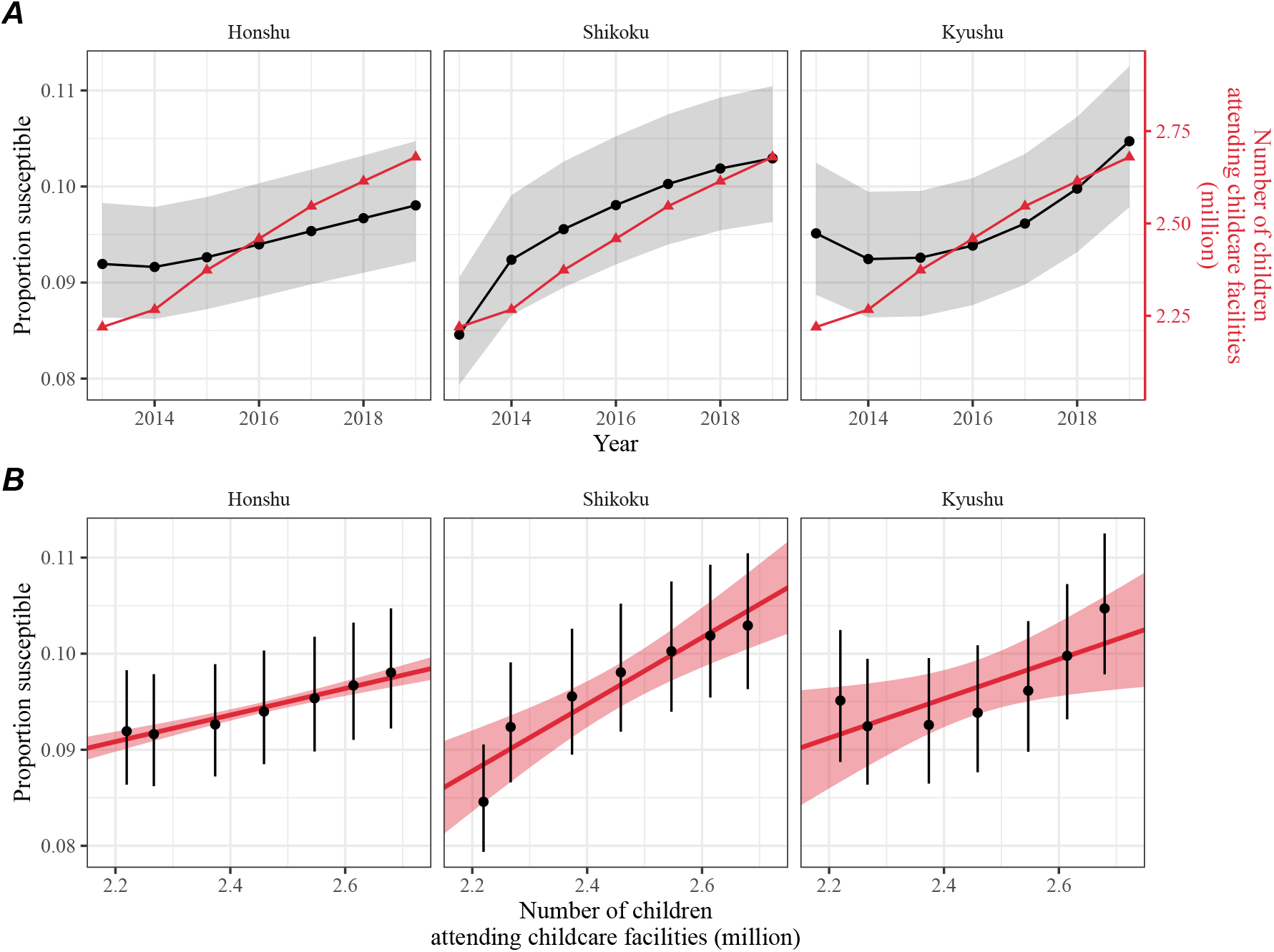
Increase in the susceptible pool following the launch of the Comprehensive Support System for Children and Childcare in Japan. (A) Direct comparisons between the estimates of susceptible proportion at the beginning of each season in each island (black) and the number of children attending childcare facilities in Japan (red). Shaded regions represent the 95% credible interval in our estimates. (B) Correlations between the estimates of susceptible proportion at the beginning of each season in each island and the number of children attending childcare facilities in Japan. Error bars represent the 95% credible interval in our estimates. Red lines and shaded regions represent the best fitting linear regression and the corresponding 95% confidence intervals.

## Discussion

We present an epidemiological analysis of RSV outbreaks in Japan combining spatiotemporal observations with dynamical disease modeling. Our analysis revealed semiannual cycles in seasonal RSV transmission in four major islands (Honshu, Shikoku, Kyushu, and Ryukyu), which correlate with specific humidity. We found that these semiannual cycles allowed a sudden shift in the seasonality of RSV outbreaks in response to an increase in population-level susceptibility. We hypothesize that an increase in childcare capacity through Comprehensive Support System for Children and Childcare may be a main driver of the increase in population-level susceptibility (DeHaan et al., 2024).

Our analysis revealed considerable heterogeneity in epidemic dynamics across major islands in Japan. We showed that these differences could be explained by the differences in underlying seasonal transmission. Notably, we found a robust, quadratic relationship between the estimated transmission rates and mean specific humidity across four major islands (Honshu, Shikoku, Kyushu, and Ryukyu), indicating low transmission at intermediate levels of specific humidity. These findings echo earlier studies that demonstrated similar relationships for RSV (Baker et al., 2019) and influenza (Lowen et al., 2007; Shaman and Kohn, 2009; Shaman et al., 2010; Tamerius et al., 2013; Lowen and Steel, 2014). The robustness of this quadratic relationship across islands exhibiting different climate conditions suggests a possibil-ity that climate-driven transmission may be, in part, facilitated by human behavior: for example, an increase in time spent indoors during low and high humidity seasons can contribute to increased transmission. We note that this relationship is correlational, rather than causal, and therefore any other climate variables (e.g., temperature or rainfall) or seasonal variation in human behavior (e.g., school terms) that correlate with seasonal variation in specific humidity will be implicitly captured by this relationship.

We tentatively hypothesized that the Comprehensive Support System for Chil-dren and Childcare may have contributed to the increase in population-level susceptibility, but other mechanisms may have contributed as well. For example, Wagatsuma et al. (2021) hypothesized that changes in climate and an increase in inbound overseas travelers may be both responsible for this shift in seasonality. While an increase in overseas travelers may also contribute to the increase in contact rates and therefore the effective susceptibility against RSV, it is likely to have weak effects given that the mean age of infection for RSV is typically very young; for example, a local surveillance effort in Kyoto reported that *>* 80% of PCR confirmed RSV infections were derived from *<* 6-year old in 2023–2024 (Matsumura et al., 2025). Another competing hypothesis that could lead to an increase in susceptibility would be strain evolution: for example, one study noted that L172Q/S173L mutant strains of RSV B that became dominant around 2016 had reduced susceptibility against monoclonal antibodies (Okabe et al., 2024). However, it is not yet clear how this mutation translates to susceptibility against infection-derived antibodies. While our findings are consistent with those by DeHaan et al. (2024), who also pointed out the association between an increase in childcare attendance and a large change in the seasonality of Kawasaki disease in Japan, we cannot rule out the possibility that other factors, such as increase in air travel (Wagatsuma et al., 2021) or strain evolution (Okabe et al., 2024), could have contributed to the increase in population-level susceptibility.

Interventions to slow the transmission of COVID-19 have disrupted the circulation of many pathogens (Baker et al., 2020; Eden et al., 2022; Chen et al., 2024; Park et al., 2024), including RSV epidemics in Japan. This disruption has added major challenges to predicting future outbreaks, which prevented us from making long-term predictions. Continued analysis of RSV dynamics in the post-COVID period, particularly with regard to whether RSV outbreaks in Japan return to fall or winter outbreaks, may help further validate our models.

Our analysis relied on several simplifying assumptions. For example, our model assumed that the waning of immunity can render previously infected individual to become fully susceptible to reinfections; this approximation allowed us to reconstruct the dynamics of susceptible hosts more easily. In practice, immunity is likely more complex with secondary infections being less susceptible and transmissible than primary infections (Pitzer et al., 2015). Other studies have also suggested the importance of interaction between RSV A and B (White et al., 2005; Holmdahl et al., 2024) as well as competition between RSV and human metapneumovirus (Bhattacharyya et al., 2015); our model did not account for such strain dynamics. We also did not account for explicit spatial structure or underlying stochasticity of the system. Therefore, our estimates of transmission rates must be interpreted with caution as they may implicitly capture factors that we did not account for explicitly, including strain dynamics, spatial structure, and exogenous transmission. Our analyses also primarily focused on three major islands (Honshu, Shikoku, and Kyushu), necessitating a better understanding of RSV dynamics in Hokkaido and Ryukyu islands; however, we note that these three islands make up *>* 95% of the population in Japan, meaning that our analyses capture the majority of RSV transmission in Japan. Despite these limitations, our model likely represents a parsimonious approximation of the complex host-pathogen interactions, allowing us to draw general conclusions about how interactions between endogenous (population-level susceptibility) and exogenous (climate-driven factors) factors can give rise to a sudden dynamical transition.

Understanding how endogenous and exogenous factors shape epidemic dynamics is critical to predicting future outbreaks and making public health decisions. Our analysis shows that the interplay between climate-driven transmission and subtle changes in population-level susceptibility can cause a sudden transition in epidemic dynamics. More broadly, our analysis demonstrates that detailed epidemiological time series data can allow us to tease apart endogenous and exogenous factors in explaining dynamical transitions, offering unique insights into a long-standing ecological question.

## Materials and methods

### Epidemiological data

The Japan prefecture-level weekly time series of RSV cases comes from the National Institute of Infectious Diseases (NIID). The NIID issues Infectious Diseases Weekly Report (IDWR) every week, which includes sentinel-reporting diseases. Specifically, RSV infections are reported through ≈ 3000 pediatric sentinel sites, which cover around 10% of pediatric institutions in Japan (Yamagami et al., 2019). We downloaded all available IDWR surveillance tables for sentinel-reporting diseases from the beginning of 2013 to end of 2023 from https://www.niid.go.jp/niid/en/survaillance-data-table-english.html and extracted RSV time series from these tables.

### Demographic data

Population sizes for each prefecture as of 2022 were obtained from Statistics of Japan website (https://www.e-stat.go.jp/en). Statistics on the number of children attending childcare facilities were obtained from the Children and Families Agency website (cfa.jp.gov).

### Climate data

The specific humidity data used in this study is from European Centre for Medium-Range Weather Forecasts (ECMWF) Reanalysis v5 (ERA5) (Hersbach et al., 2020). The original data are hourly with a horizontal resolution of about 31km. We first resample the hourly data to obtain daily mean values and then perform spatial average over cell grids within each prefecture in Japan. We further summarized the daily time series of specific humidity into weekly mean values in each prefecture, which were further averaged over to obtain weekly mean values in each island.

### Center of gravity and epidemic trough

In order to characterize changes in the timing of the epidemic, we quantified the center of gravity of RSV cases for each RSV season at each prefecture. Here, we excluded the Okinawa prefecture, which is the southernmost prefecture of Japan, due to differences in RSV seasonality: in contrast to all other prefectures that exhibit winter outbreaks, summer outbreaks are observed in the Okinawa prefecture. To compute the center of gravity, we defined the RSV season from week 27 of the current year to week 26 of the next year and numbered each week of season from 1 (starting from week 27 of a given year) to 52 (ending at week 26 of the following year); this simplification allows us to track changes in RSV seasonality in a consistent manner. Then, for each season, we calculated center of gravity by taking the weighted mean of the week of season, weighted by the number of cases. We added 26 to the resulting center of gravity to convert the estimates to be in the units of regular weeks (rather than the week of season). For each season, we also quantified the corresponding epidemic trough by taking the minimum value of weekly cases. For the 2019–2020 season, we took the minimum cases before 2020 to exclude the impact of COVID-19 interventions.

### Transmission and observation model

To model the population-level spread of RSV in Japan, we extended the standard Susceptible-Infected-Recovered-Susceptible (SIRS) model to account for non-sinusoidal seasonal transmission rates and changes in transmission patterns due to COVID-19 intervention measures. Specifically, the discrete-time SIRS model is given by:

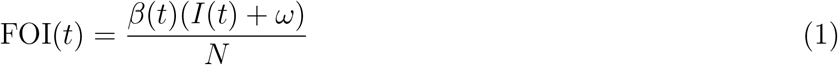

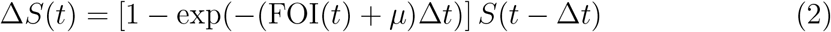

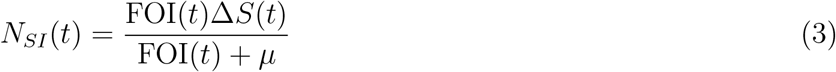

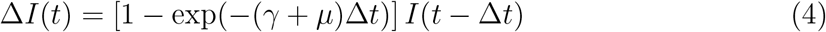

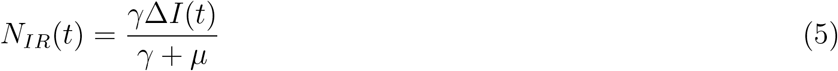

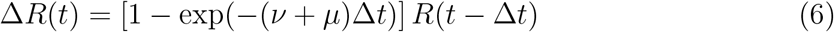

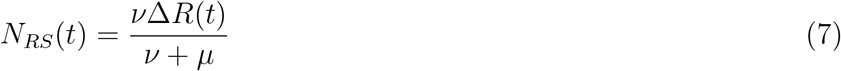

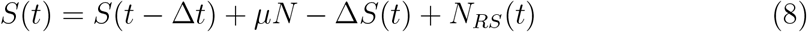

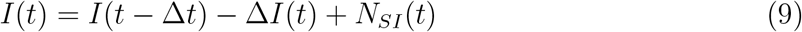

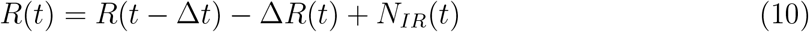

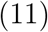

Here, *S, I*, and *R* represent the number of individuals who are susceptible, infected, and recovered; *N* represents the total population size; FOI(*t*) represents the force of infection at time *t*; Δ*X*(*t*) represents number of individuals who leave the compartment *X* at time *t*; *N*_*XY*_ (*t*) represents the number of individuals who move from compartment *X* to compartment *Y* at time *t*; *β*(*t*) represents the time-varying transmission rate; *ω* represents the number of imported infections; *γ* represents the recovery rate; *ν* represents the immune waning rate; and *µ* represents the birth and death rates. This model assumes a simple demography and extreme waning, which allows immune individuals to become fully susceptible. Therefore, model parameters must be interpreted with care, especially the duration of immunity. In practice, re-infection can still occur even under partial immunity, in which case the duration of immunity can be shorter (Pitzer et al., 2015).

Typically, the transmission rate is assumed to follow a sinusoidal function for modeling endemic diseases. Instead, we decomposed *β*(*t*) into a product of two separate terms:

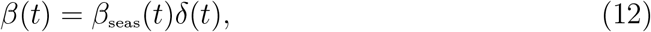

where *β*_seas_(*t*) represents the seasonal transmission rate and *δ*(*t*) represents relative changes in transmission due to COVID-19 intervention measures, such that *δ <* 1 represents transmission reduction. A similar decomposition was recently used for modeling the spread of Mycoplasma pneumoniae infections (Park et al., 2024).

First, we modeled the seasonal transmission rate *β*_seas_(*t*) as a periodic function with a period of 52 weeks (*β*_seas_(*t*) = *β*_seas_(*t −* 52)) and tried to estimate a separate value for each week. To constrain the shape of *β*_seas_(*t*), we imposed cyclic random-walk priors:

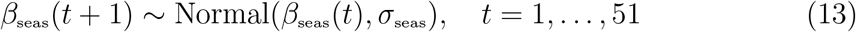

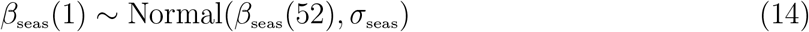

where the standard deviation *σ*_seas_ determines the smoothness of the seasonal transmission rate. We imposed a weakly informative prior on *σ*_seas_:

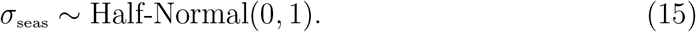

To further constrain the range of seasonal transmission rate *β*_seas_(*t*), we imposed additional priors:

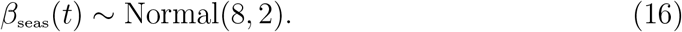

Second, we assumed *δ*(*t*) = 1 for *t <* 2020 (before the COVID-19 pandemic) and tried to estimate a separate value for *δ*(*t*) at each week. To constrain the shape of *δ*(*t*), we imposed random-walk priors:

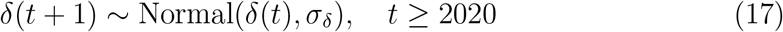

where the standard deviation *σ*_seas_ determines the smoothness of the estimated *δ*(*t*). We imposed a weakly informative prior on *σ*_*δ*_:

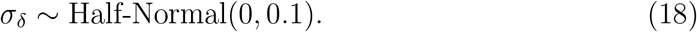

To further constrain the range of estimated intervention effects *δ*, we imposed additional priors:

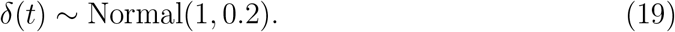

For the analysis of Hokkaido island, we estimated *δ*(*t*) beginning from 2017 instead of 2020 to capture the sudden transition to irregular epidemic dynamics occurred in 2017; we tried fitting a model that estimated *δ*(*t*) beginning from 2020 but found that it was unable to explain the sudden transition. For the analysis of Ryukyu island, we estimated *δ*(*t*) beginning from 2019 instead of 2020 to capture the unusually large RSV outbreak that happened in 2019.

We assumed that the recovery rate *γ* = 1*/*week and birth/death rates *µ* = 1*/*(80 × 52) weeks are known. We imposed weakly informative priors on all other parameters:

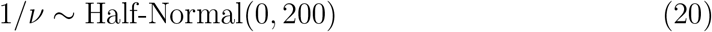

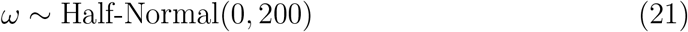

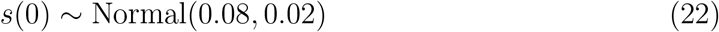

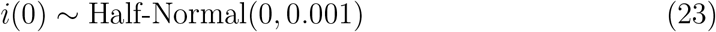

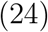

where *s*(0) and *i*(0) represent the initial proportion of susceptible and infected individuals such that the initial conditions are given by: *S*(0) = *Ns*(0) and *I*(0) = *Ni*(0).

Finally, the model was fitted to case data in each island by assuming a negative binomial observation error:

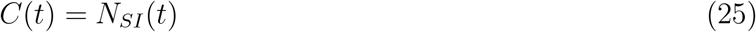

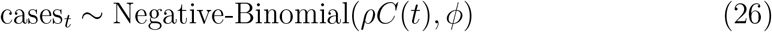

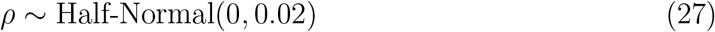

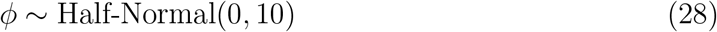

where *C*(*t*) represents the incidence of infection, *ρ* represents the under-reporting rate, and *ϕ* represents the overdispersion parameter. Parameter estimation was performed in a Bayesian framework using the rstan package (Carpenter et al., 2017). Convergence was assess by ensuring low R-hat, high effective sample size, no divergent transitions, and no iterations that exceeded the maximum tree depth. The model struggled to converge for the analysis of Shikoku island—in this case, removing the Normal(1, 0.2) prior on *δ*(*t*) allowed us to fit the model.

### Simulations

We run a series of simulations to understand how the interplay between climate-driven transmission and population-level susceptibility can drive a sudden shift in the timing of an epidemic. First, we simulated the model for a year (from week 26 of the starting year to week 25 of the following year) by varying the initial conditions and computing the center of gravity. Specifically, we varied *i*(0) between 1 × 10^*−*4^ and 3 × 10^*−*3^ and *s*(0) between 0.078 and 0.105. All other parameters were set to posterior median estimates.

To further understand how the shape of seasonal transmission term affects the degree of shift in seasonality, we varied the shape of seasonal transmission term by interpolating the estimated *β*_seas_ from Honshu and Kyshu islands, which are the two most populated islands. To do so, we first took posterior median estimates of *β*_seas_ from two islands and fitted generalized additive model with cyclic cubic spline bases to obtain smoothed estimates of *β*_seas_ for each island, which we denote as *β*_*H*_ and *β*_*K*_, respectively. Then, we normalized seasonal transmission rates such that it has a mean of zero and has an amplitude of 1:

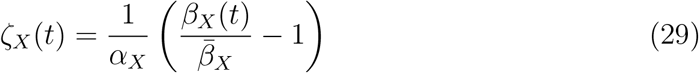

where *ζ*_*X*_(*t*) represents the normalized seasonal transmission pattern in island *X*, and *α*_*X*_ = (max(*β*_*X*_(*t*)) − min(*β*_*X*_(*t*)))*/*2 represents the amplitude of seasonal transmission pattern in island *X*. This allowed us to interpolate between two normalized seasonal terms and obtain a flexible shape for seasonal transmission rate:

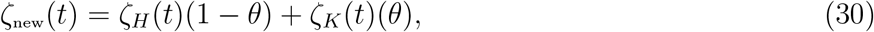

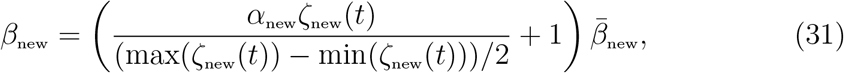

where *θ* represents the interpolation coefficient, such that *θ* = 0 and *θ* = 1 causes *β*_new_ to have the same shape as *β*_*H*_ and *β*_*K*_, respectively and 0 *< β <* 1 allows us to model counterfactual transmission scenarios that interpolates between two islands. Note that *ζ*_new_(*t*) does not necessarily have an amplitude 1 so we divide it by (max(*ζ*_new_(*t*)) *−* min(*ζ*_new_(*t*)))*/*2 to ensure the amplitude of 1.

For a given value of the interpolation coefficient *θ* and seasonal amplitude *α*_new_, we simulated two outbreaks for a year assuming *s*(0) = 0.078 and *s*(0) = 0.105 and computed the difference in the timing of epidemic peak. In doing so, all other parameters, including the mean transmission rate *β*_new_, were fixed to posterior median estimates for the Honshu island.

### Regression

We also performed a linear regression between the estimated proportion of susceptibles at the beginning of each season (week 26) against the number of children attending childcare facilities. For simplicity, the regression was performed using median estimates for the susceptible proportions. We did not have data on the number of children attending childcare facilities broken down by island level and so we used the national-level data instead.

## Data availability

All data and code are stored in a publicly available GitHub repository (https://github.com/parksw3/perturbation).

## Acknowledgements

This project has been funded in whole or in part with Federal funds from the National Cancer Institute, National Institutes of Health, under Prime Contract No. 75N91019D00024, Task Order No. 75N91023F00016. The content of this publication does not necessarily reflect the views or policies of the Department of Health and Human Services, nor does mention of trade names, commercial products or organizations imply endorsement by the U.S. Government. S.W.P. is a Peter and Carmen Lucia Buck Foundation Awardee of the Life Sciences Research Foundation. I.H. received postdoctoral funding from the High Meadows Environmental Institute of Princeton University. B.T.G. and C.J.E.M. acknowledge support from Princeton Catalysis Initiative and Princeton Precision Health.

## Supplementary Materials

### Supplementary Figures

**Figure S1:**
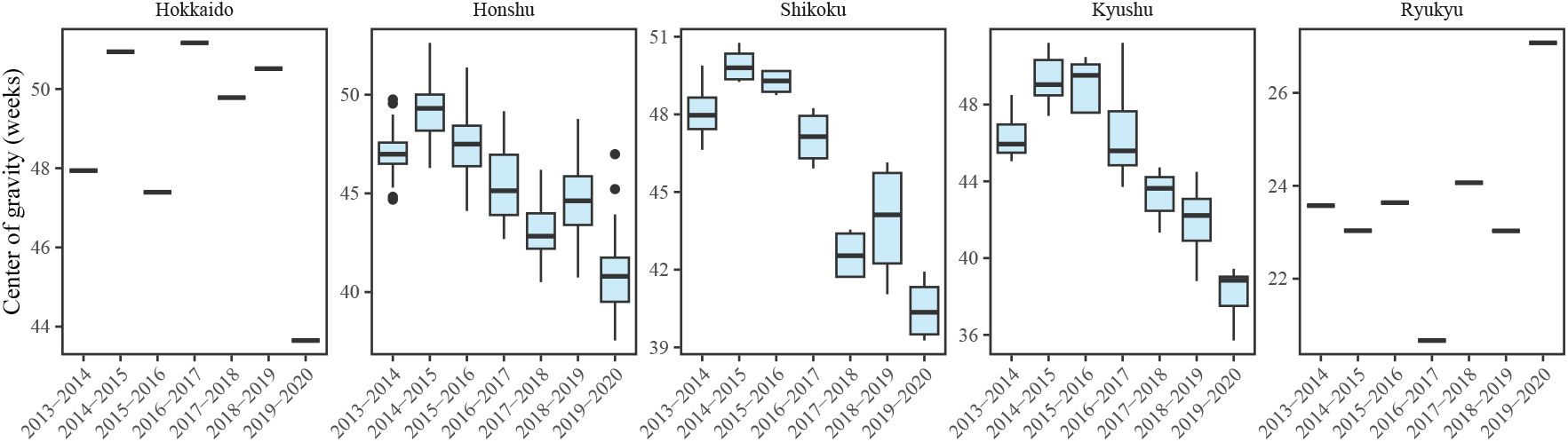
Estimates of center of gravity (i.e., the mean timing of an epidemic) across all prefectures, stratified by island. Hokkaido and Ryukyu islands each contain only one prefectures: Hokkaido and Okinawa, respectively.

**Figure S2:**
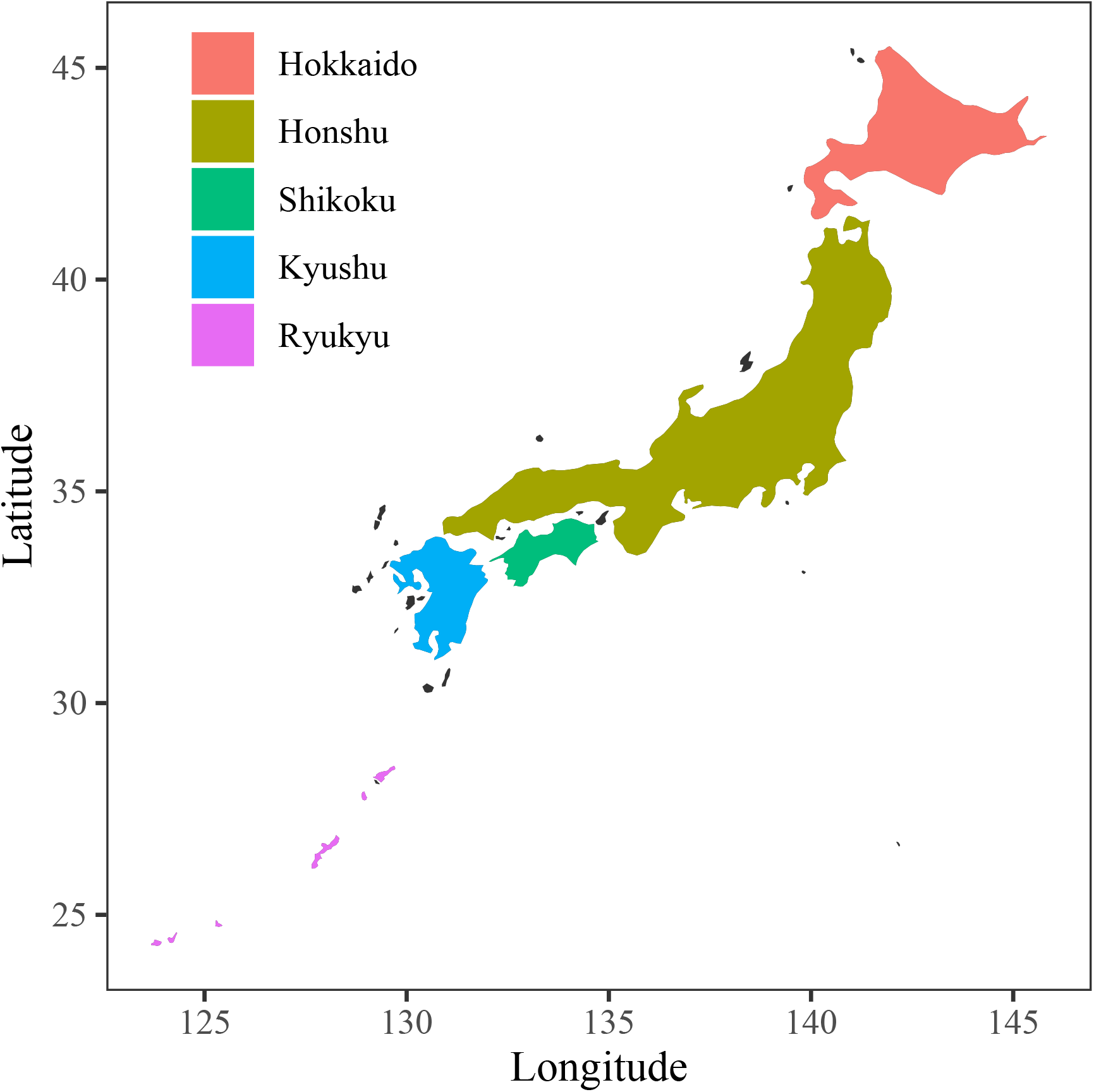
Colored map of Japan. Each of five major islands are marked by different colors.

**Figure S3:**
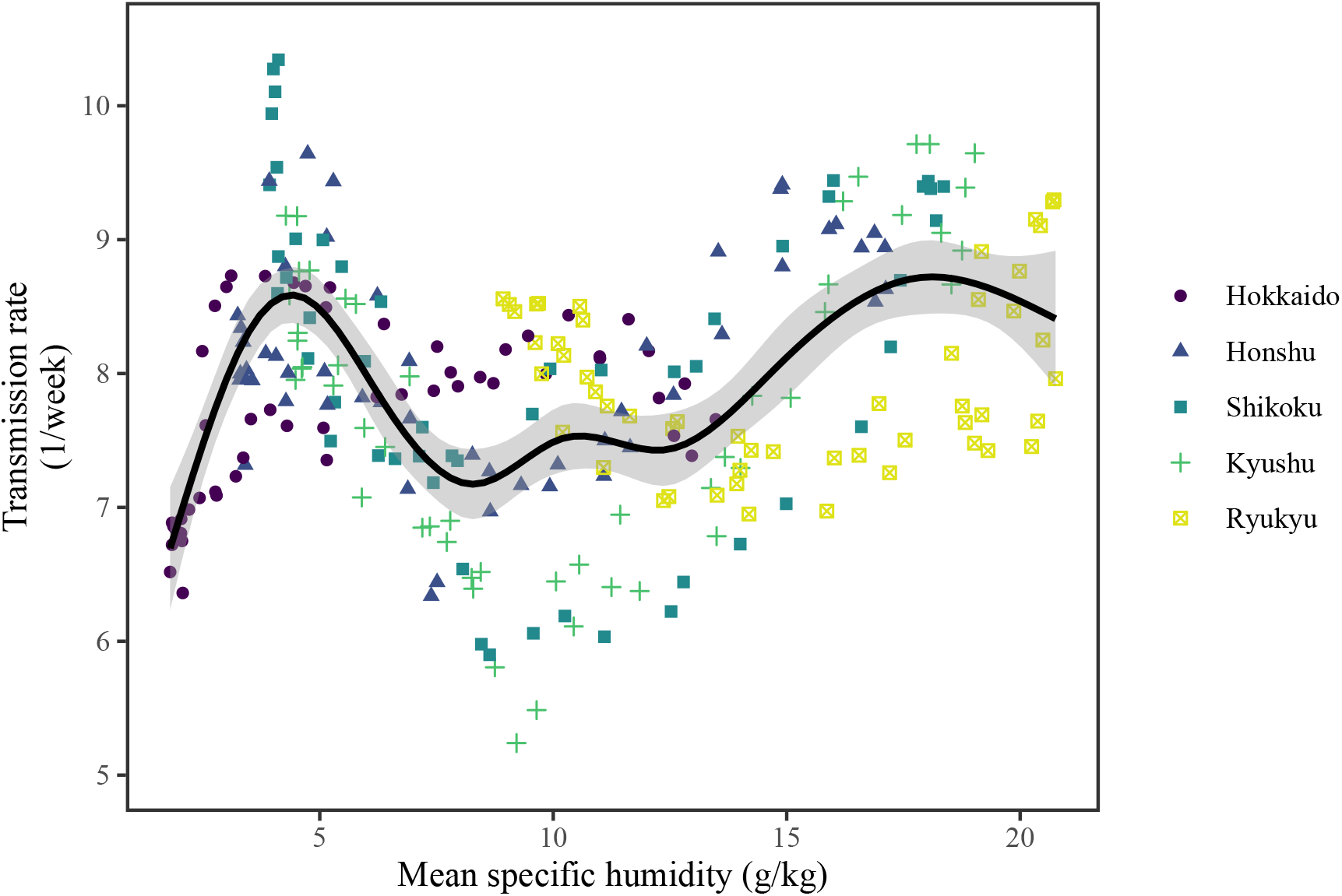
Joint relationship between the estimated periodic seasonal transmission rates and mean specific humidity across all five islands. Points represent the estimates across 52 weeks in each island. The line represent a generalized additive model fit using cubic spline basis.

**Figure S4:**
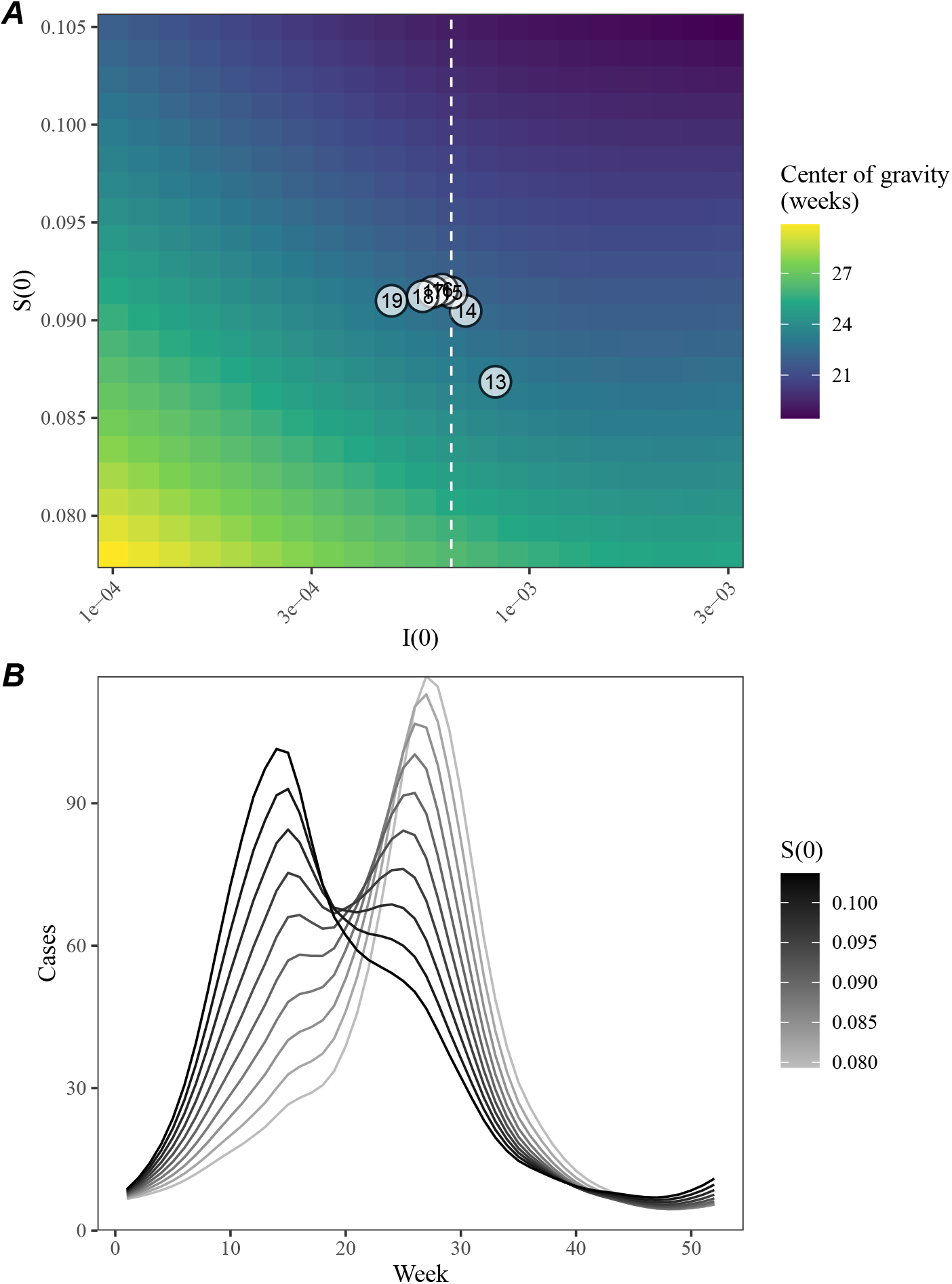
A lack of increase in susceptible pool explains constant seasonality. (A) Predicted effects of the proportion of infected *i*(0) and susceptible *S*(0) at the beginning of season on center of gravity. Points represent the estimated values for *i*(0) and *s*(0) between 2013 and 2019, showing the last two digits of a given year. The white vertical dashed line represents the *i*(0) value used for simulating epidemic dynamics in panel B. (B) Changes in epidemic trajectories that would be caused by an increase in the susceptible proportion at the beginning of season for a fixed value of *i*(0).

## Notes

### Competing Interest Statement

The authors have declared no competing interest.

